# An AluYb8 retrotransposon characterises a risk haplotype of TMEM106B associated in neurodegeneration

**DOI:** 10.1101/2023.07.16.23292721

**Authors:** Alex Salazar, Niccolò Tesi, Lydian Knoop, Yolande Pijnenburg, Sven van der Lee, Sanduni Wijesekera, Jana Krizova, Mikko Hiltunen, Markus Damme, Leonard Petrucelli, Marcel Reinders, Marc Hulsman, Henne Holstege

**Affiliations:** Section Genomics of Neurodegenerative Diseases and Aging, Department of Clinical Genetics, Vrije Universiteit Amsterdam, Amsterdam UMC, Amsterdam, The Netherlands; Delft Bioinformatics Lab, Delft University of Technology, Delft, The Netherlands; Alzheimer Center Amsterdam, Neurology, Vrije Universiteit Amsterdam, Amsterdam UMC location VUmc, Amsterdam, The Netherlands; Amsterdam Neuroscience, Neurodegeneration, Amsterdam, The Netherlands; Institute of Biomedicine, University of Eastern Finland. Kuopio, Finland; Institute of Biochemistry, Christian-Albrechts-University Kiel, Otto-Hahn-Platz 9, Kiel, Germany; Department of Neuroscience, Mayo Clinic, Jacksonville, FL, USA; Neuroscience Graduate Program, Mayo Clinic Graduate School of Biomedical Sciences, Jacksonville, FL, USA

## Abstract

Genome-wide association studies identified a role for *TMEM106B* in various neurodegenerative diseases. Based on long-read whole-genome sequencing data of 256 individuals, we identified an AluYb8 retrotransposon in the 3’ UTR of the risk haplotype of *TMEM106B*. When transcriptionally active, Alu-elements can propogate throughout the genome, and mediate (post-)transcriptional dysregulation of nearby genes. We found that *TMEM106B* haplotypes carrying the AluYb8 element are more methylated than those without, likely reflecting an evolutionary selection to suppress propagation. AluYb8 activation can be further suppressed by TDP-43, in its role in post-transcriptional RNA-processing. However, age-related loss of TDP-43, by reduced methylation in the 3’ UTR of *TARDBP,* may release AluYb8 suppression. Together, our findings suggest that in the aging brain, the AluYb8 insertion may mediate dysregulation of *TMEM106B*, impacting the endolysosomal system via a negative-feedback loop, ultimately leading to neurodegenerative disease. Notably, *TMEM106B* haplotype sequences are different between African and European genomes, which likely explains the different effects on disease-risk between both populations. Overall, our research advances the understanding of the roles of TDP-43 and TMEM106B in neurodegenerative diseases, and provides a novel connection between genetic variation and age-related changes in genomic and cellular regulation.

## Introduction

*TMEM106B* likely plays a central role in neurodegenerative diseases. The gene encodes a 274 amino-acid transmembrane protein highly expressed in the central nervous system, especially in neurons and oligodendrocytes^1,2^. Although its functions are not entirely clear, TMEM106B localizes in the endolysosomal system, where it is involved in maintaining lysosome integrity ^1,2^. Genome-wide association studies (GWAS) have highlighted the presence of two common haplotypes in *TMEM106B* (allele-frequency ∼50%), one associated with increased risk of neurodegenerative diseases, and hence the other with a protective effect^2–10^ (see Figure 1 A). The haplotypes are characterised by several common variants that are in strong-to-complete linkage disequilibrium (LD) with each other (see Figure 1 A and B). The rs1990622 variant was associated with protection against frontotemporal lobar degeneration with TAR DNA binding protein inclusions (FTLD-TDP) (OR=0.61; p = 1.08×10^-11^)^3^, which may extend to amytrophic lateral sclerosis (ALS)^11^. The rs13237518 variant was associated with protection against Alzheimer’s disease (OR=0.96, p = 4.90×10^-11^)^12^. Notably, the association of the protective haplotype in *TMEM106B* with FTDLD-TDP is more pronounced in carriers of pathogenic *GRN* variants (OR=0.49, p = 1.34×10^-9^)^2,3^, a gene encoding progranulin (PGRN)^13^. Since both genes are essential for lysosome function^13^, their effects may converge in the impairment of lysosomal mechanisms.

**Figure 1.**
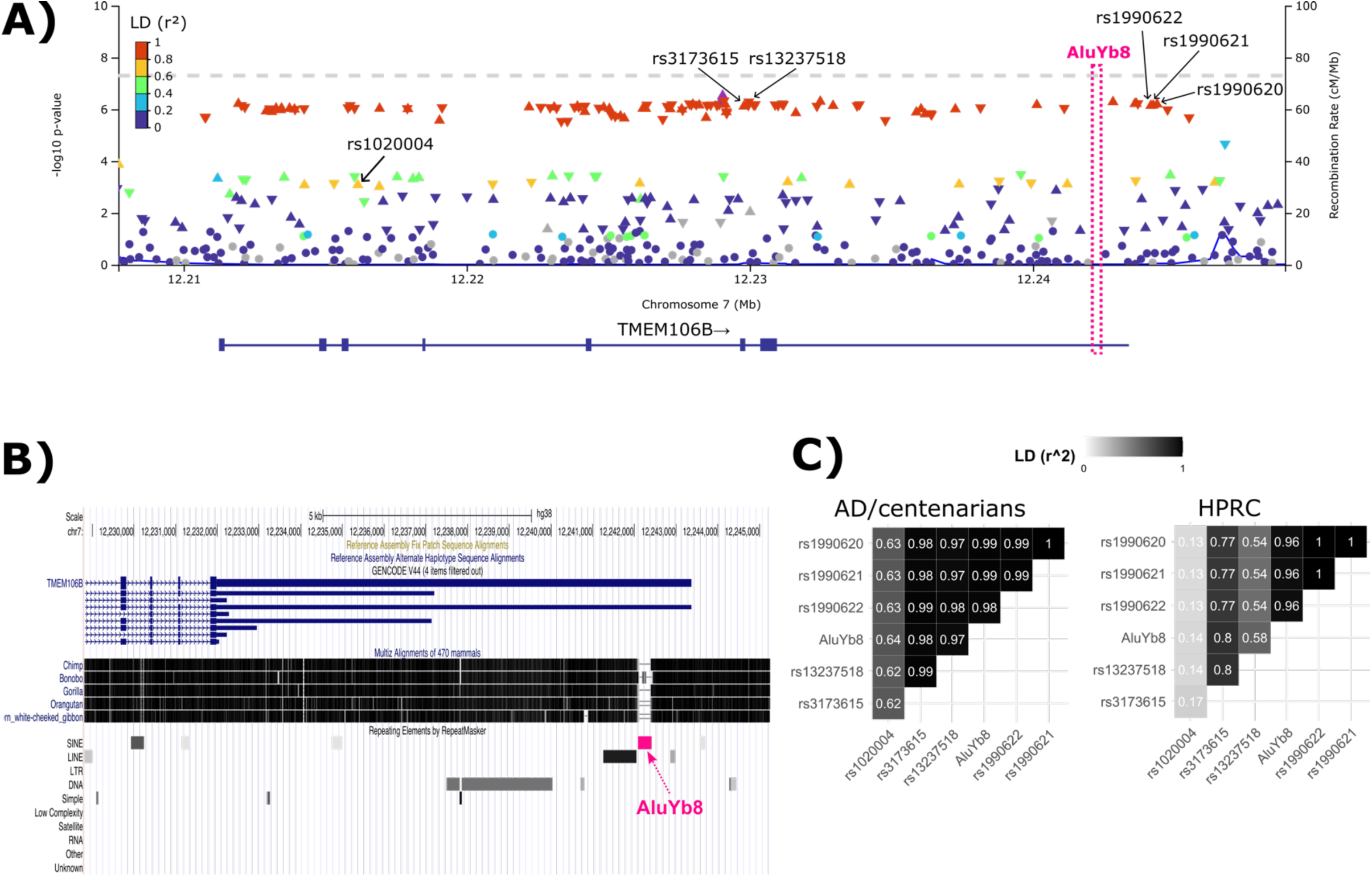
An AluYb8 retrotransposon further characterizes risk and protective haplotypes of TMEM106B associated in neurodegeneration. Modified overview of the TMEM106B gene from LocusZoom^86^ displaying genome-wide associated (GWAS) variants (triangles) from the Bellenguez *et al.* 2022 study^12^; the color of each variant represents LD (r^2^) values based on genomes with European ancestry. **(B)** Screenshot from the UCSC genome browser displaying: (top) the last 3 exons along with the 3’ UTR of known transcripts from *TMEM106B* (blue rectangles); (middle) whole-genome alignments to great-ape reference assemblies; and (bottom) repeat annotations with the location of the AluYb8 element highlighted in pink. Note that the AluYb8 is human-specific as it is absent in the great ape reference genomes. **(C)** Pairwise LD (r^2^) values based on 92 AD genomes and 117 centenarian genomes from The Netherlands^4,80^ (left), and 47 genomes from Human Pangenome Reference Consortium (HPRC)^28^ (right). LD values between *TMEM106B* risk alleles and rs1020004 is expectedly reduced as previously reported^12^.

Both the rs1990622 and the rs13237518 variants are in strong linkage with rs3173615, a non-synonymous variant (T185S) which substitutes a threonine residue (risk) for a serine residue (protective) in exon 6 of *TMEM106B*^2,9^. This substitution has become the focus of many studies aiming to explore the functional consequences of the protective haplotype of *TMEM106B*^2,5,14–18^. The T185S substitution is predicted to be ‘benign’ by Polyphen, SIFT and REVEL (score of 0.0267), suggesting that the subsitution is unlikely to have major effect on TMEM106B function. The T185S substitution juxtaposes an *N-*glycosylation site at position 183, which has been proposed to possibly affect post-translational protein maturation^19^. However, *N-*glycosylation sites commonly occur at a N-X-S/T motif, suggesting that substituting the N_183_-I_184_-T_185_ motif to N_183_-I_184_-S_185_ motif is unlikely to have any effect^19^. Notably, there were no observable functional changes in a CRISPR-based knockin-mouse model homozygous for the T185S substitution^14^.

Nevertheless, several studies hypothesized that T185S may alter *N-*glycosylation such that the risk haplotype has a higher aggregation potential^18–24^. Aggregate amyloid fibrils composed of TMEM106B protein were observed in the post-mortem brain of humans with advanced age, or brains from individuals suffering from neurodegenerative diseases^20–24^. They were specifically composed of the C-terminal fragment (CTF) of TMEM106B^20–24^, and carriers of the risk haplotype had higher amounts of CTF compared to carriers of the protective haplotype^18^. Cryogenic-electron microscopy showed that the aggregated fibrils have three distinct structural polymorphisms: fold I is predominant in individuals homozygous for the risk allele threonine (TT) and heterozygous carriers (TS); fold II represents only a small fraction of TMEM106B CTF fibrils in carriers of the risk haplotype; and fold III is predominant in individuals homozygous for the protective allele serine (SS).^20–24^. This suggests that fold III may have lower aggregation potential than folds I and II. Note that there is currently no consensus that either haplotype leads to increased or decreased gene expression levels or TMEM106B protein abundance^2,5,8,10,14,15,17,25,26^. Another variant —rs1990620, which is in complete LD with the protective haplotype^10^—reportedly affects long-range chromatin interactions leading to decreased recruitment of CCCTC-binding factors (CTCF), potentially influencing *TMEM106B* transcription levels^10^. However, multi-tissue eQTL analysis of rs1990622 using the UKBEC database shows no association^5^, while a similar analysis of the protective rs1990622 and rs3173615 using the GTEx database indicates an association with *increased TMEM106B* expression in the cerebellum, cortex, and cerebellar hemisphere^5^.

Thus far, studies have exclusively focused on single nucleotide variants from GWAS and/or short-read whole-genome sequencing data. Here, we explored whether a 317 bp insertion, previously reported to be in linkage with the risk haplotype of *TMEM106B*^27^, may be the driver of the genetic association with altered disease risk.

## Results

### An AluYb8 element further differentiates the risk and protective haplotypes of TMEM106B

In a PacBio whole-genome long-read sequencing dataset of genomes from 92 Dutch Alzheimer Disease cases, 117 Dutch centenarians and 47 individuals from diverse human populations from HPRC (Human Pangenome Reference Consortium^28^), we observed a common ∼317 bp insertion in the 3’ untranslated region (UTR) in the risk haplotype of *TMEM106B* which is ∼12.7 Kbp downstream of rs3173615 (see Table 1, see Figure 1A). The total length of this insertion varied from 306-330 bp with a median sequence length of 317 bp, largely due to length variation of a poly-A tail in the 3’-end of the sequence. Annotations from the GRCh38 reference genome, along with sequence alignment to the DFAM repeat database^29^, showed that the insertion is an Alu retrotransposon of the Yb8 lineage sub-family (see Figure 1B). Alu-elements are retrotransposons that comprise ∼11% of all human genomic sequences^30–32^. They propagate through a “copy-and-paste” mechanism by using reverse-transcription of their mRNA transcripts to then insert themselves at different regions in the genome^30–32^. Importantly, Alu-elements are primate-specific; the Yb8-lineage is among the youngest lineage of Alu-elements (integrated ∼2.39 million years ago), and more dominantly expanded in humans than other primates^30,33,34^. Indeed, the AluYb8 element is human-specific as it is absent in the great ape reference genomes (see Figure 1B).

**Table 1.**
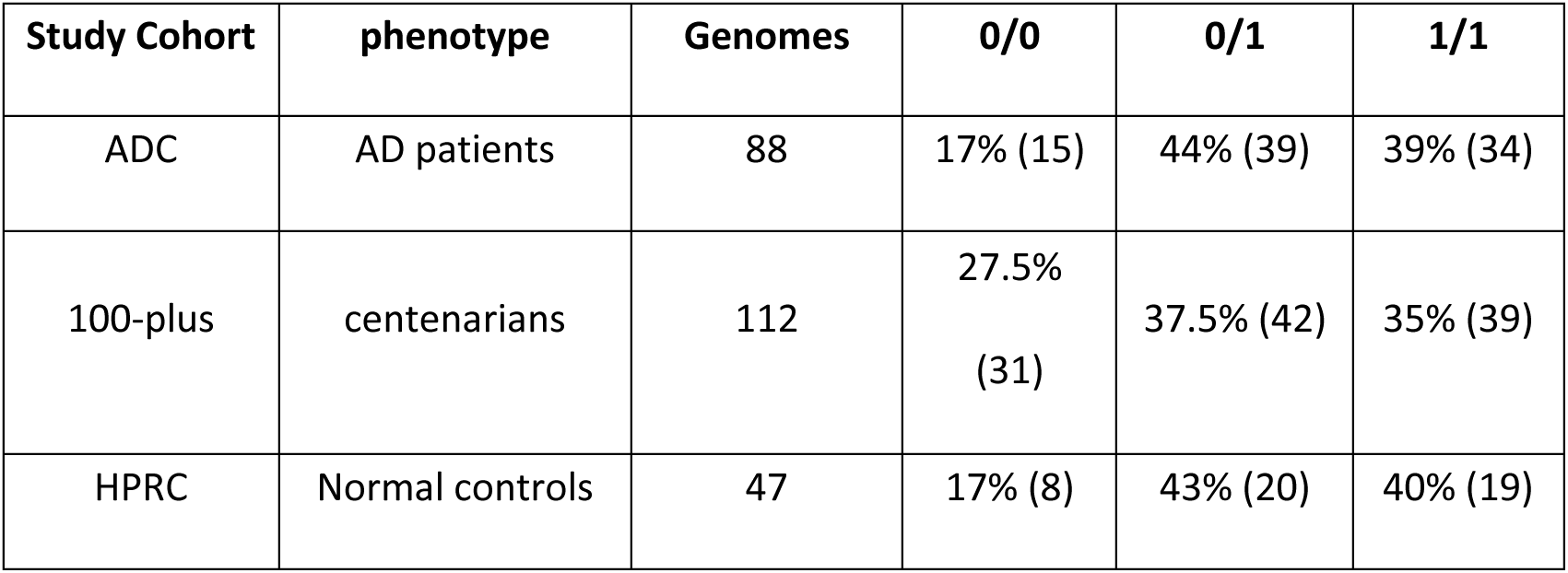
An AluYb8 retrotransposon is commonly present in the 3’ UTR of TMEM106B. Observed relative frequencies of homozygous non-carriers (0/0), heterozygous (0/1), and homozygous carriers (1/1) of the AluYb8 element in long-read sequencing datasets of Alzheimer’s disease (AD) and cognitively healthy centenarian genomes from The Netherlands (100-plus study), as well as genetically diverse genomes from the Human Pangenome Research Consortium (HPRC).

### Epigenetic differences between the TMEM106B haplotypes may yield functional consequences

Transcriptionally active retrotransposons can dysregulate nearby gene expression and translation^31,35^, hence, they are often supressed by the host through DNA methylation^36^. Therefore, we investigated potential differential methylation status of CpG islands between the *TMEM106B* haplotypes. Using the single-molecule methylation signals captured during PacBio HiFi whole-genome sequencing of the Dutch AD and centenarian genomes, we identified a total of 231 unique CpG methylation sites in *TMEM106B* (see Figure 2A). From these CpG sites, 39 were significantly differentially methylated in homozygous carriers the AluYb8 element relative to homozygous non-carriers (Wilcoxon rank-sum test, p-val ≤ 1.43×10^-4^, see Figure 2A and 2B); with 27 sites having at least a 10% higher methylation (mean increase of 63%) in carriers of the risk haplotype (see Figure 2A and 2B). Most of these 27 sites affected intronic regions in *TMEM106B*, including relatively older (retro)transposable elements such as ancestral L1 sequences (L1PA6, L1PA7, and L1ME3G), the J and S Alu-based sub-families (AluSg and AluJb), and the TcMar-Tigger family (Tigger1). Additionally, eight sites that affected the 3’ UTR of *TMEM106B* (outside the AlubY8 element insertion), had at least a 10% higher methylation level in carriers of the risk haplotype compared to carriers of the protective haplotype (Figure 2A and 2B). Notably, two CpG sites with increased methylation in the risk haplotype affected two separate enhancer sequences: one site with a 15% methylation increase immediately downstream of exon 1 (accession code EH38E2534496); and a second site with a 50% methylation increase ∼1,500 bp upstream of exon 4 (accession code EH38E2534502), which encodes for the amino acid residues defining the C-terminal fraction (CTF) of TMEM106B protein found in the aggregated fibrils of neurodegenerative and aged human brains^20–24^ (see Figure 2A). The possible consequence of these differential methylation levels on the expression of the CTF remains unclear.

**Figure 2.**
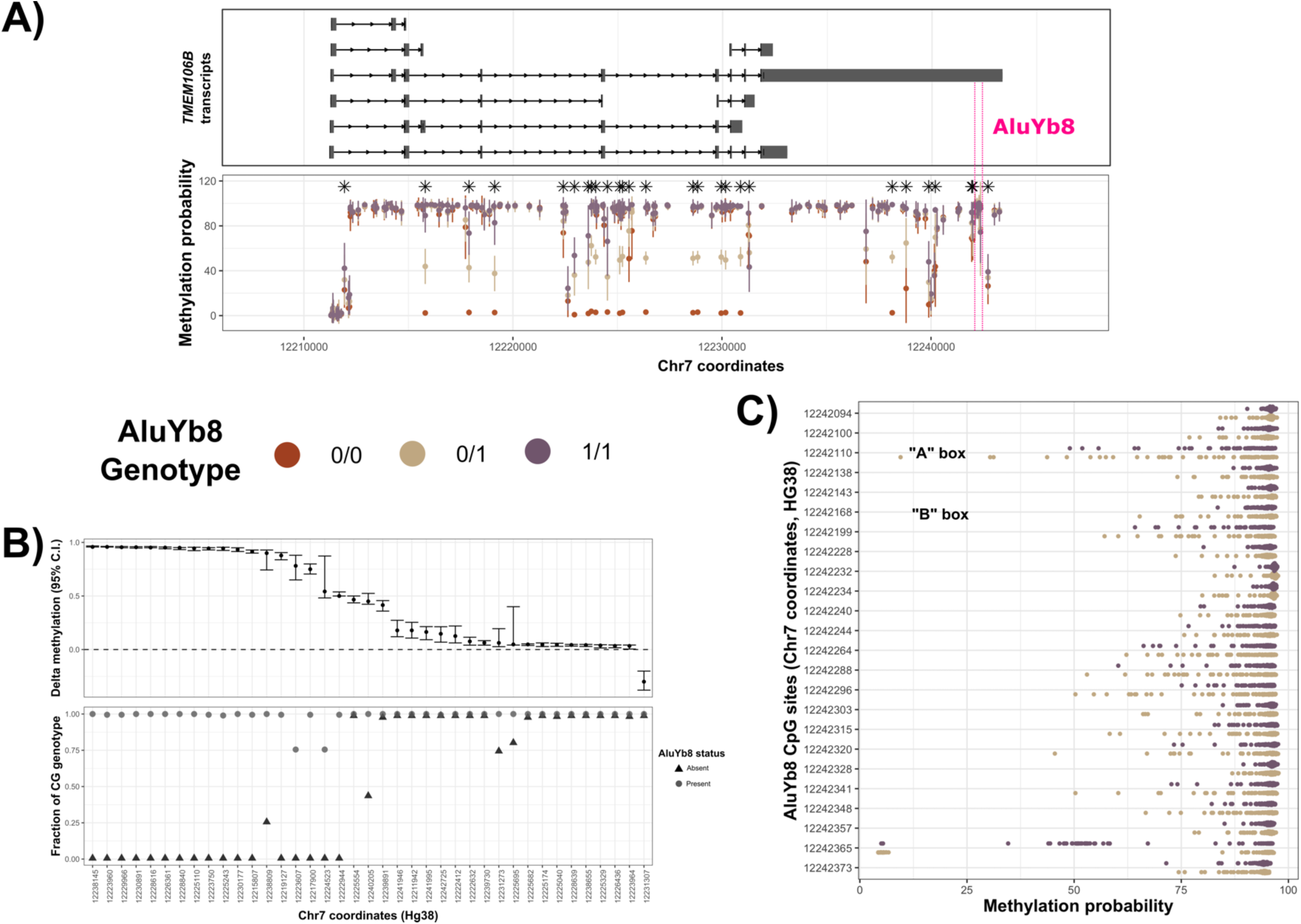
The risk and protective haplotype of *TMEM106B* vary in their epigenetic landscapes. Distribution of methylated CpG sites in *TMEM106B* based on PacBio HiFi whole-genome sequencing of peripheral blood from AD and centenarians. **(A)** *TMEM106B* transcripts from *Ensembl*^85^ (top), overlayed with relative coordinates of each CpG site (bottom), displaying the average methylation probability (dots) along with the corresponding standard deviations (vertical bars). Asterisks indicate significantly different CpG sites from (**B)**. **(B)** 39 CpG sites were found to be significantly different between homozygous carriers (1/1) and homozygous non-carriers (0/0) (Wilcoxon rank-sum test, p-value ≤ 2.42×10^-4^, Bonferroni corrected). Y-axis shows the estimated methylation difference between carriers and non-carriers with the 95% confidence interval, while X-axis shows the coordinates of the CpG sites in the GRCh38 reference genome sorted by descending estimated difference. CpG sites in carriers are generally more methylated. **(C)** Methylation probability distributions (Y-axis) for each CpG site inside the AluYb8 element (X-axis) between heterozygous and homozygous AluYb8 carriers.

Only one of the 39 CpG sites was significantly *less* methylated in carriers of the risk haplotype compared to carriers of the protective haplotype. We observed a 27% methylation decrease within an MIRb sequence—an ancient transposable element in mammalian genomes^37–39^ (Figure 2A and 2B). Notably, MIRb sequences were found to be enriched in human long-noncoding RNA molecules (lncRNAs) that are localised in the nucleus^39^, suggesting gene and chromatin regulatory properties. Hence, demethylation may influence retention of MIRb in lncRNA transcripts, potentially modulating (protective) regulatory effects in *cis-* and/or *trans-*.

Of the 39 differentially methylated CpG sites, 19 sites can be explained by single nucleotide variants present in the risk haplotype but absent in the protective haplotype (see Figure 2B), likely reflecting an evolutionary selection for CpG sites to suppress retrotransposon activity. This is particular true for CpG sites with more than a 50% methylation difference between homozygous carriers of the risk and protective haplotypes (see Figure 2B), as unique cytosine-guanine dinucleotides in the risk haplotype enables methyation, which is otherwise absent in the protective haplotype.

The majority of the CpG sites inside the AluYb8 sequence were methylated across all individuals that carry the risk haplotype, reflecting active suppression of the retrotransposon^36^ (see Figure 2C). However, methylation levels varied widely between individuals (see Figure 2C), suggesting inter-individual variation in potential AluYb8 activation. Two specific CpG sites in the AluYb8 element had the highest variability, one of which is inside the “A box” RNA polymerase III recognition sequence^40^ (see Figure 2C). Demethylation of the “A” and “B” box recognition sequences have been shown to increase transcription of Alu elements^34,41,42^. We also observed methylation variability of an additional CpG site in the 3’ end of the AluYb8 sequence (see Figure 2D), largely driven by a SNP in a subset of risk haplotype sequences (AF = 0.236), changing a CG dinucleotide to CA. The CG allele is reportedly part of a conserved 3’ UTR stem-loop in a subset of active Alu-elements^43^. Although this SNP likely does not perturb Alu transcription, it is unclear if it decreases retrotransposition, as it may disrupt binding recognition from critical enzymatic machinery required for downstream DNA integration^43,44^. We did not observe significant methylation differences between AD and centenarian genomes (although power was limited in this small sample).

### The 3’ UTR of TARDBP is variably methylated in peripheral blood of AD and centenarian individuals

*TARDBP* encodes for the TDP-43 protein, which with its role in post-translational RNA processing, can also suppresses retrotransposon transcription. However, TDP-43 commonly aggregates in aged and neurodegenerative human brains^35,45,46^. The 3‘ UTR of *TARDBP* is reportedly demethylated with age, consequently dysregulating transcription and translation, which reduces the activity of TDP-43^47^. As a consequence, demethylation of *TARDBP* may lead to increased Alu-element expression^36^, connecting TDP-43 dysregulation with the dysregulation of *TMEM106B* transcribed by the risk allele^31,35^. We thus investigated the methylation status of *TARDBP* in context with the AluYb8 element insertion in the risk haplotype of *TMEM106B*.

Overall, we found 255 unique CpG sites in the *TARDBP* gene across AD and centenarians, with an average of 240 total CpG sites per genome. We found no significant differences in CpG methylation per site between AD and centenarians, although statistical power is limited (see Figure 3A). In the 3’ UTR, we found 31 unique CpG sites for which methylation status was highly variable between individuals, especially towards the end of the 3’ UTR (see Figure 3B). This finding is in agreement with Koike *et al.* 2021, who reported that gene expression and alternative splicing of *TARDBP* can be dysregulated via age-related demethylation of a specific subregion in the 3’ UTR harbouring a cluster of 15 CpG sites^47^ (see black bar on top of Figure 3B). In particular, demethylation of the six left-most CpG sites was found to suppress alternative splicing and increase *TARDBP* gene expression^47^ (Figure 4C). In our genomes, we identified 14/15 of these CpG sites: the six left-most sites showed similar demethylation patterns as those observed in controls and ALS patients from Koike *et al.* ^47^ (see Figure 3B and 3C). These findings suggest that age-related demethylation of the 3’ UTR of *TARDBP* also occurs in peripheral blood, which may lead to dysregulation of genes regulated by Alu-elements.

**Figure 3.**
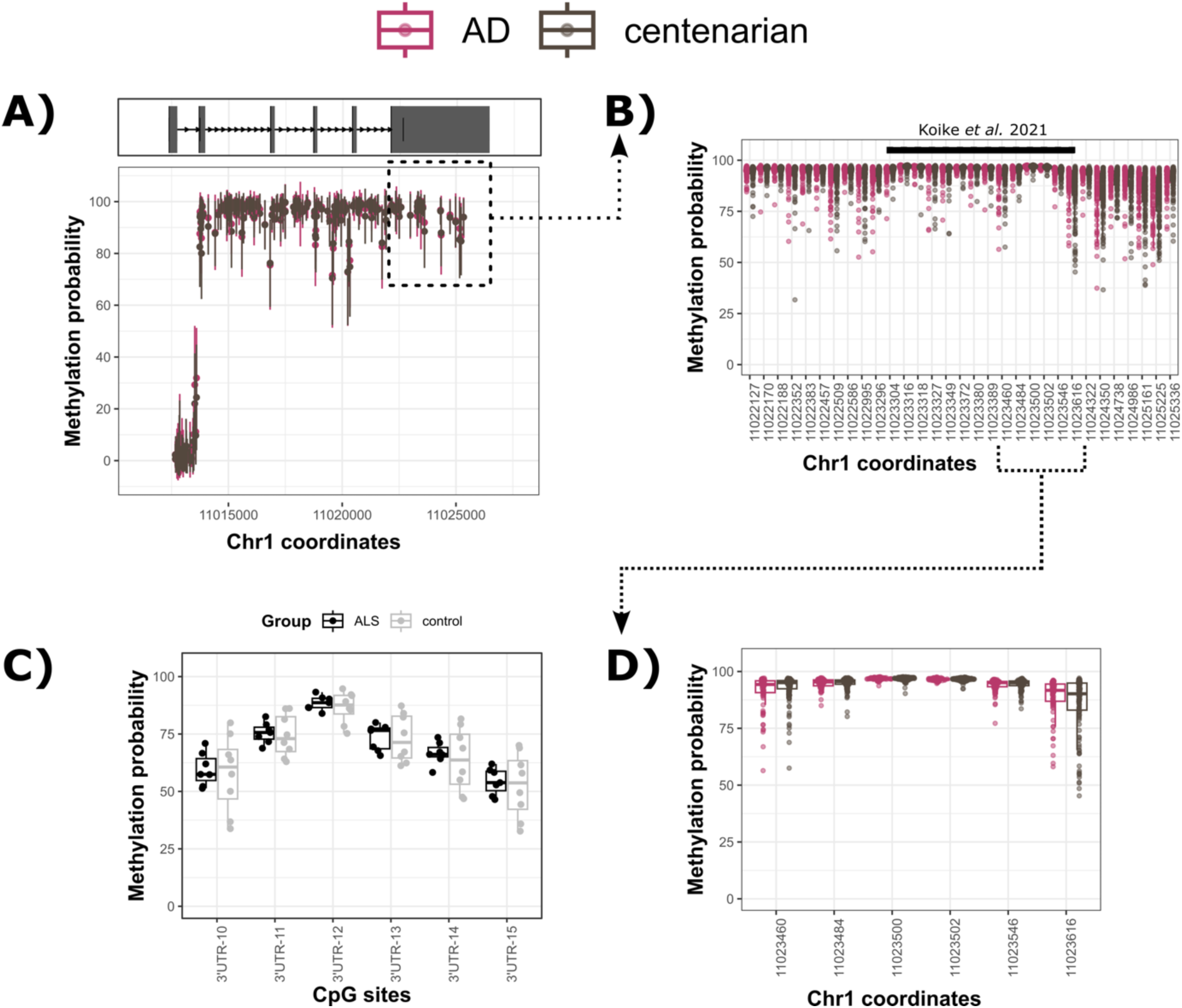
The 3’ UTR of *TARDBP* is variably methylated. Distribution of methylated CpG sites in *TARDBP* based on PacBio HiFi whole-genome sequencing of peripheral blood from AD and centenarians. **(A)** Canonical transcript of *TARDBP* based on *Ensembl*^85^ annotations (top), with the corresponding methylation status of CpG sites found within *TARDBP* region between AD (violet) and centenarian (dark grey) individuals (bottom). **(B)** Same as (A), but displaying only CpG sites within the 3’ UTR of *TARDBP*. The black horizontal bar on top depicts CpG cluster in *TARDBP* reported by Koike *et al*. 2021^47^, where demethylation of **(C, D)** the six left-most CpG sites can dysregulate *TARDBP* transcription. **(C)** Displays methylation data for the six left-most CpG sites from motor neurons from amyotrophic lateral sclerosis (ALS) patients and control individuals as reported by Koike *et al*. 2021^47^. **(D)** Same as (C), but from peripheral blood from AD and centenarian individuals from this study.

**Figure 4.**
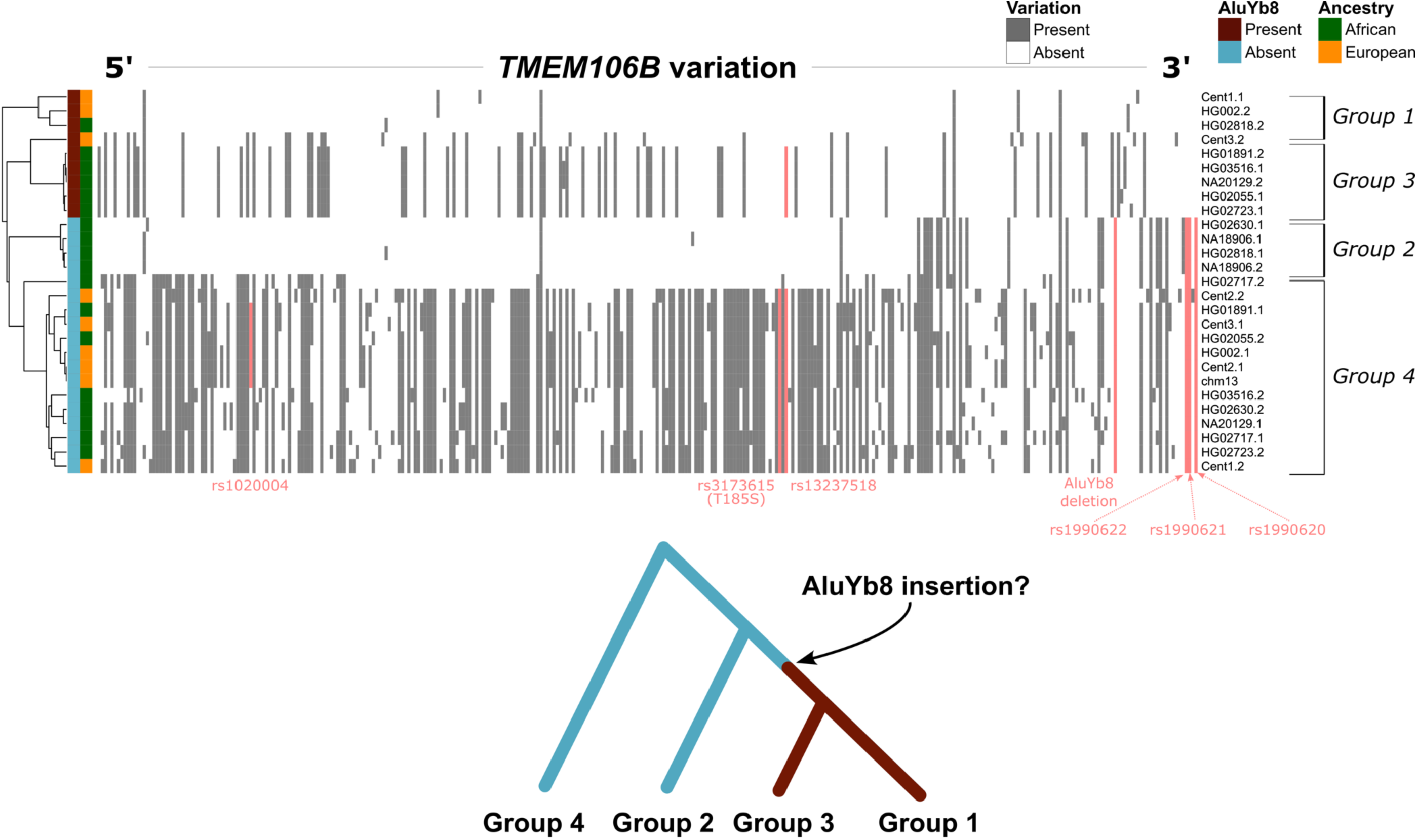
Comparison of phased *de novo* assemblies reveals novel haplotypes in *TMEM106B* in genomes with African ancestry. (Top) Single-nucleotide and (large) structural variants were identified in 27 complete phased sequences of *TMEM106B* from 14 long-read *de novo* genome assemblies, where CHM13, HG002, and Cent1-3 are from European ancestry (orange); the rest are genomes from African ancestry with reduced linkage disequilibrium (dark green, see Figure 1C). Each row represents one of two haplotype sequences (.1 or .2 suffix) coloured by presence (maroon) or absence (blue) of the AluYb8 element in the 3’ UTR. Columns represent all variation jointly identified in all *TMEM106B* haplotype sequences based on whole-genome alignment to the GRCh38 reference genome, ordered from their relative position from 5’ to 3’ orientation; grey bars represent the binary presence of each variant in each haplotype sequence, with selected variants previously associated with neurodegeneration (see Figure 1) are highlighted (salmon colour) along with the deletion of AluYb8. Clustering of the binary presence/absence of all variation yields four major clusters, annotated as haplotype groups 1 to 4. **(Bottom)** *Speculative* evolutionary history of the *TMEM106B* haplotype sequences.

### Convergent evolution of SINE-element insertions in the 3’ UTR of TMEM106B in non-primate species may offer downstream survival advantage

Previous studies have highlighted a potential evolutionary convergence for selection of SINE retrotranposition insertion in the 3’ UTR of various genes in human and rodent genes^48,49^, suggesting that such events can alter local regulatory capacity, and provide a survival advantage through additional regulation of biological pathways. Indeed, for the *TMEM106B* gene, we found that species-specific SINE retrotransposons independently inserted in the 3’ UTR of *TMEM106B* in other mammals. In the *Mus musculus* reference genome (GRCm39), we observed an insertion of a “B-element” from the B4 family—a a rodent-specific SINE retrotransposon^50^—in the orthologous 3’ UTR of *TMEM106B*. Sequence analysis showed that the insertion site similarly involved a AT-rich sequences including an A-based homopolymer, reflecting previously observed bias of SINE insertions in AT-rich regions of 3’ UTRs^48^. Long-read whole-genome sequencing from 20 genetically distinct mice^51^ indicated that they were all homozygous for the B4-element. Likewise, the rat (*Rattus norvegicus*) reference genome (RGSC 6.0) also harbours the B4-element in orthologous 3’ UTR of *TMEM106B*, suggesting that this retrotransposon predates the evolutionary split between mouse and rat. Interestingly, the pig reference genome (Sscrofa11.1) also harbours an insertion of a PRE0_SS retrotransposon in an AT-rich site of the 3’ UTR of *TMEM106B*, which is a recent SINE element of PRE1-family^52^. Genomes from modern dogs and cats (Carinivora order) are an exception with regard to retrotransposons in the 3’UTR in TMEM106B: while these Carnivora-specific Can-SINEs elements exist^53,54^, we did not observe them in the 3’ UTRs of their respective TMEM106B homologs.

### Long-read phasing of TMEM106B reveals novel haplotypes in African ancestry

In the analyzed Dutch genomes, the AluYb8 element is in complete LD with the reported variants characterising the risk haplotype of *TMEM106B* (r^2^ = 0.98 – 1.0, p < 2.2×10^-16^; Figure 1C). In the HPRC dataset, we also observe complete LD between the AluYb8 and the downstream SNPs, but the LD between AluYb8 element and the upstream risk variants is lower (r^2^=0.5-1.0, p < 2.2×10^-16)^) (Fig 1C). We found that this reduced LD was driven by 9 genomes with African ancestry: HG02630 (Gambian subpopulation), HG02717 (Gambian), HG02818 (Gambian), HG01891 (African Caribbean in Barbados), HG02055 (African Caribbean in Barbados), HG02723 (Gambian), HG03516 (Esan in Nigeria), NA18906 (Nigeria), and NA20129 (African Ancestry in Southwest USA). We hypothesised that this difference may be due to a (meiotic) recombination event(s) in the region between rs13237518 and the AluYb8, which may have given rise to an additional haplotype(s) in the *TMEM106B* locus, unique for Africans and their descendants.

To further characterize the different haplotypes in *TMEM106B,* we compared haplotype-phased *de novo* assemblies between the African genomes with reduced LD and the following European genomes: HG002 (Eastern European, Jewish-Ashkenazi ancestry), CHM13 (European ancestry^55^) genomes, and three Dutch centenarian genomes (Cent1, Cent2, and Cent3; see Figure 4). We found a total of 341 unique variants across *TMEM106B* sequences, and binary clustering revealed four major clusters (see Figure 4). Two haplotype sequence clusters—termed groups 2 and 3—were composed mostly of African genomes. The AluYb8 element was absent in groups 2 and 4, and present in groups 1 and 3. Notably, risk alleles in the 5’-end and the 3’-end of *TMEM106B* are separated in haplotypes groups 2 and 3. More specifically, group 2 harbours 1990622, rs1990621, and rs1990620 risk alleles, but not rs3173615 (T185S) and rs13237518. In contrast, group 3 only harbours the rs13237518 risk allele without rs3173615 (T185S). In short, compared to genomes from European ancestry, genomes from African ancestry carry alternate haplotype sequences in the *TMEM106B* locus, defined by a breakpoint between rs13237518 and the AluYb8 element (see Figure 1C and 4).

## Discussion

Long-read sequencing revealed that the risk haplotype of the *TMEM106B* locus includes an AluYb8 retrotransposon in the canonical transcript of the 3’ UTR of *TMEM106B* gene. Alu-based insertions are known to (dys)regulate transcription and translation of nearby genes^31,56,57^, making the AluYb8 element a candidate driver of the neurodegenerative effect associated with the *TMEM106B* risk haplotype. We further observed that CpG islands in the risk and protective haplotypes were differentially methylated, potentially influencing *TMEM106B* transcription due to differential binding affinity of RNA polymerases and regulatory elements. TDP-43 has previously been shown to suppress retrotransposon transcription, including Alu-elements^35,45,46^. We found that the 3’ UTR of *TARDBP*, where age-related demethylation reportedly dysregulates TDP-43^47^, is variably demethylated in the peripheral blood of AD patients and in centenarians. Our results lead us to propose a mechanism in which the presence of the AluYb8 element in *TMEM106B*—along with age, TDP-43 leads to increased risk for neurodegeneration via a negative feedback loop, mediated by the AluYb8 element. We hypothesize that pathogenic PGRN variants may provide an additive negative effect on this feedback loop (Figure 5).

**Figure 5.**
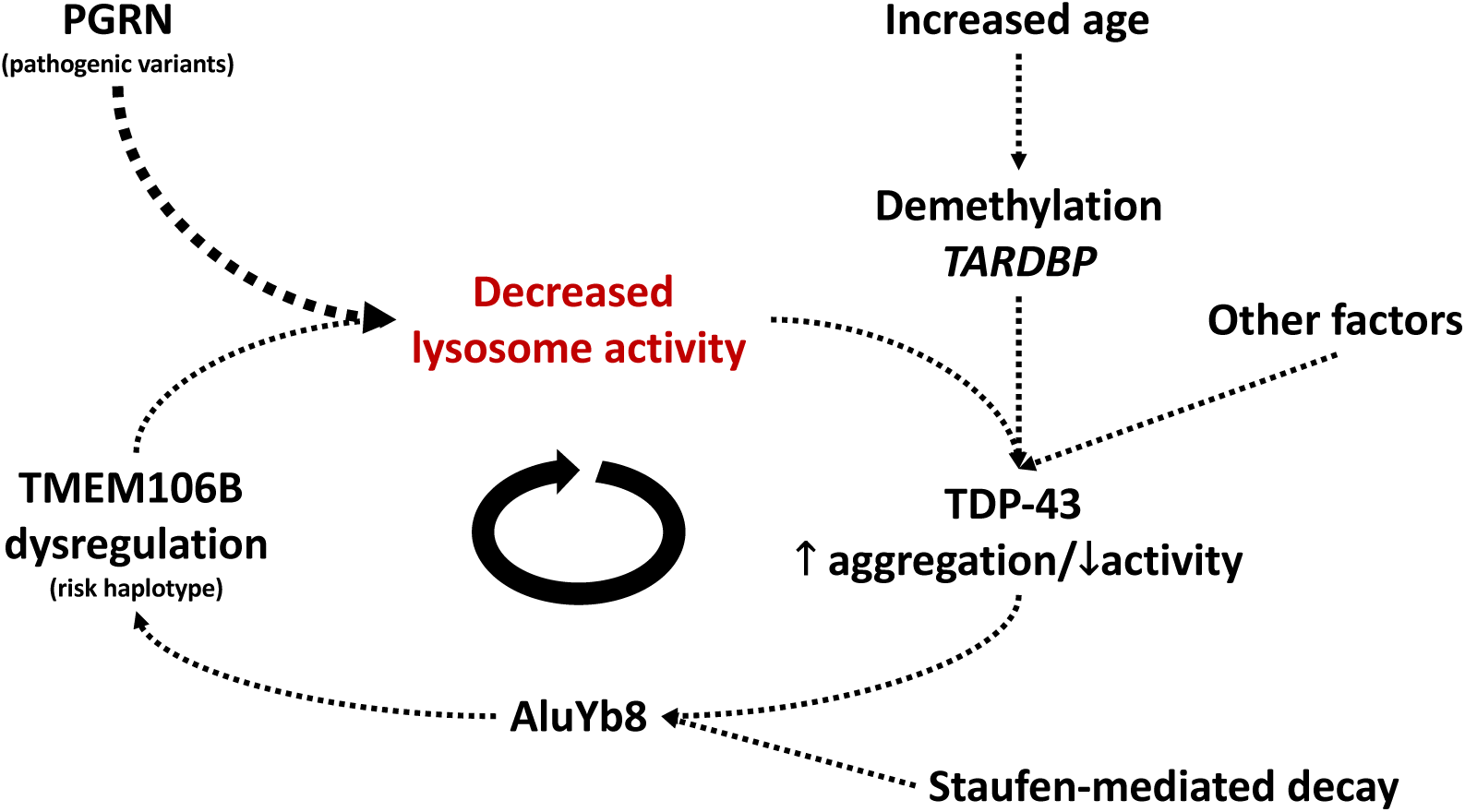
Endolysosomal processes depend on GRN, TMEM106B, TDP-43 and advanced age interactions. Pathogenic *GRN* variants leads to compromised endolysosomal activity, in turn increasing aggregation of TDP-43 due to incomplete degradation. Increased age leads to hypomethylation of *TARDP* (gene encoding TDP-43), which dysregulates TDP-43 protein synthesis. As TDP-43 supresses Alu-element transcription, reduced functionality of TDP-43 leads to increased Alu-element transcription, consequently dysregulating TMEM106B and further compromising endolysosome activity.

A transcriptionally active AluYb8 can propagate throughout the host genome, leading to genomic instability^36^. Hence, loci with Alu-insertions are under evolutionary selection pressure to recruit CpG sites that are commonly hypermethylated by their host to suppress Alu-element activation^36^. Indeed, we found that the *TMEM106B* risk haplotype had higher methylation levels than the protective haplotype. However, aging commonly leads to a global decrease in methylation in humans^58,59^, which may consequently lead to an age-related increased expression of transposable elements^60–62^. Under this scenario, RNA polymerase III (which transcribes Alu-elements^63^) is thought to compete with RNA polymerase II (which transcribes genes) at overlapping regions along the DNA^35,64^, ultimately leading to decreased gene expression and decreased protein abundance of nearby genes. This may be further excarberated via better binding of RNA III due to demethylation of the “A” box promoter sequence in the AluYb8 element. Additionally, the transcriptional suppression of retrotransposons as mediated by the RNA-processing activity of TDP43 may be decreased with age, due to demethylation of 3’ UTR of *TARDBP*^35,45,46^. In this study, we observed the demethylation of both the “A” promoter sequence and the 3’ UTR of *TARDBP*.

The presence of a AluYb8 element may further mediate *TMEM106B* dysregulation, even when it is in a transcriptionally inactive state. For example, long non-coding RNAs (lncRNAs) and micro-RNA (miRNAs) in *cis-* and *trans-* can bind to transcripts containing Alu elements^65–67^, leading to mRNA degradation via Staufen-mediated decay^67,68^. Indeed, miRNAs have been previously shown to bind to the 3’ UTR of *TMEM106B*, leading to transcriptional suppression^16^. Moreover, impaired splicing due to loss of TDP-43 activity may further exacerbate degredation of *TMEM106B* transcripts. Finally, *cis-* and *trans-* interactions between the AluYb8 and lnc- and mi-RNAs, methylation status, and other regulatory elements may lead to a differential chromatin state^69,70^, potentially leading to altenative splicing.

Recent (functional) studies support the above mechanisms. First, (brain) tissue-specific eQTL and pQTL databases highlight that the risk haplotype of *TMEM106B* leads to lower gene expression and protein abundance than the protective haplotype^27^. Furthermore, *in vitro* TDP-43 knockdown experiments in cell lines with the *TMEM106B* risk haplotype lead to significant decrease of TMEM106B expression levels compared to cell lines with the protective *TMEM106B* haplotype^71,72^. Recent studies report that the level of the C-terminal fragment of TMEM106B (CTFT) increases in carriers of the risk haplotype in aged and neurodegenerative brains^73^. One explanation for this may be that the risk haplotype impairs splicing^74^, leading to aggregation of C-terminal segments of TMEM106B. Overall, these observations suggest that the risk haplotype of TMEM106B associates with an array of potential complex interactions involving age, TDP-43 activity and impaired endolysomal activity.

Sequence analysis of non-primate mammals genomes showed that species-specific SINE retrotransposons have independently inserted in 3’ UTR of *TMEM106B* orthologs, highlighting evolutionary convergence for selection for SINE-like regulatory elements in *TMEM106B*. This suggests that the regulatory utility of the SINE elements may provide a downstream survival advantage^48^. Staufen-mediated decay guided via SINE-retronsposon elements in 3’ UTR of genes may have convergently evolved in non-primate genomes, suggesting evolutionary significance^48,49^. Indeed, SARS-CoV-2 was shown to use TMEM106B as an alternative entry receptor for infection in human lung cells^75,76^, and increased *TMEM106B* expression correlates with SARS-CoV-2 infection^75,76^. Hence, it is possible that downregulation of TMEM106B may minimise viral infections in certain tissues (i.e. lung), but leads to a decrease in the endolysosomal activity in brain over age. Although in non-primates, SINE-like retrotransposons may have similar effects on survival as the human Alu element, it is unclear whether they have similar effects on neurodegenerative mechanisms. Certain interactions with age-related factors, such as TDP43 signaling and activity, may be human-specific. Hence, caution should be taken when using animal models to elucidate associated regulatory processes.

Lastly, a comparison of complete haplotype sequences between European and African genomes revealed four major haplotype groups in the *TMEM106B* locus, two of which are dominantly defined by African individuals, explaining why linkage disequilibrium (LD) of the risk alleles is reduced in *TMEM106B* in genomes with African ancestry. Therefore, next to the proposed mechanism of the *TMEM106B* risk haplotype, this study also highlights how the lack of complete haplotype characterisations in other ancestral populations leads to a limited understanding of neurodegenerative risk loci in non-Europeans^77,78^. Ongoing efforts to generate long read sequencing data from diverse population-backgrounds^28^ will enable rigorous evaluation of the evolutionary development of disease-associated haplotypes, and ultimately for population specific disease risk-assessments. For *TMEM106B*, we cannot exclude that additional haplotypes in the human population exist, nor that the additional haplotype groups observed in this study are entirely unique to Africans.

## Conclusion

An AluYb8 retrotransposon residing in the 3’UTR of *TMEM106B* is commonly present in the human population, and is in complete linkage with the risk haplotype of *TMEM106B* in genomes with European ancestry. Convergent evolution in non-primate genomes suggests that the *TMEM106B* risk haplotype may associate with increased survival, possibly by mitigating viral infections in lung tissue. However, our findings suggest that in the aging brain, the risk haplotype may mediate a negative feedback-loop involving the interaction of TDP43 and TMEM106B, ultimately leading to neurodegeneration by reduced endolysosomal activity.

## Methods

### Sample populations

The 92 Alzheimer disease (AD) patients in this study are from the Amsterdam Dementia Cohort (ADC) and were diagnosed as previously described^79^. The 117 cognitively healthy centenarians in this study are individuals from the 100-plus Study^80^. The Medical Ethics Committee of the Amsterdam University Medical Center approved all studies. All participants and/or their legal representatives provided written informed consent for participation in clinical and genetic studies.

### PacBio HiFi long-read whole-genome sequencing

Genomic DNA of AD and centenarian individuals were extracted from peripheral blood, and gDNA integrity was assessed with Qubit system 2 (Qubit dsDNA HS Assay Ref. No. Q32854) and Nanodrop (NanoDrop 2000). Specifically, we required DNA to have a purity of 260/230 of 2.00 and above, and 260/280 of 1.8 and above. When requirements were not met, a clean-up step was performed with 20 minutes incubation at room temperature with 1:1 AMPure PB beads (Pacific Biosciences Ref. No. 100-265-900) and two times washing with 80% ethanol. DNA is eluted with 120 µl Elution buffer (Pacific Biosciences Ref. No. 101-633-500). gDNA was sheared with the Megaruptor from Diagenode with a long hydropore cartridge (Cat. No. E07010003), using insert concentration of 116 µg with a shearing speed of first 30, and afterwards 31. Qubit dsDNA HS Assay (Ref. No. Q32854) was used to quantify DNA concentration. DNA damage repair was performed using SMRTbell Enzyme Clean Up Kit 2.0 (Ref. No. 101-932-600) with the reaction volumes as described in the manufacterers instructions. More specifically, a nuclease treatment on the formed SMRTbell library was done by preparing a master mix with SMRTbell Enzyme Clean Up Mix (Pacific Biosciences Ref. No.101-932-900), SMRTbell Enzyme Clean Up Buffer 2.0 (Pacific Biosciences Ref. No. 101-932-700) and Molecular biology Grade water. Reaction mix was added to the library complex and incubated at 37°C for 30 minutes. SMRTbell Library was purified using 1.x AMPurePB beads (Pacific Biosciences Ref. No. 100-265-900) and two times washing with 80% ethanol, library complexes were eluted in 31 µl Elution buffer (Pacific Biosciences Ref. No. 101-633-500).

All libraries were prepared using either SMRTbell Template Prep Kit 2.0 (Pacific Biosciences Ref. No. 100-938-900) or 3.0 (Ref. no. 102-141-70) following manufacturer’s instructions. Size selection was performed using the BluePippin high-pass DNA size selection (sage Science BLU0001; 0.75% Agarose Cassettes, with Marker S1). The range selection mode was set from 8,000-50,000 bp. Qubit dsDNA HS assay was used to quantify DNA concentration. Libraries having the desired size distributions were identified on the Femto pulse System (Agilent M5330AA) using the 48kb Ladder. Fractions centered between 15 kb, and 21 kb were used for sequencing.

Polymerase binding for all sequencing libraries were prepared using Sequel® II Binding Kit 2.2 (Pacific Biosciences Ref. No. 101-894-200) and 3.2 (Pacific Biosciences Ref. No. 102-194-100) following manufacturer’s instructions. Sequencing primers were conditioned by heating to 80°C for 2 minutes and rapidly cooled to 4°C. The sequencing primer was annealed to the template at a molar ratio calculated by PacBio for 15 minutes at room temperature. After primer annealing, polymerase was bound to the primed template for 15 minutes at room temperature. Polymerase-bound samples were then kept at 4°C prior to use. Prior to sequencing, excess unbound polymerase was removed by incubating the complexes for 5 minutes with 1.2× (vol:vol) AMPure PB beads (Pacific Biosciences Ref. No. 100-265-900) at room temperature. Beads were not washed with 80% ethanol. After removing the free polymerase in the supernatant, the polymerase-bound complexes were eluted with Adaptive loading buffer (Pacific Biosciences Ref. No. 102-030-300). Sequencing reaction was performed using the Sequel sequencing kit 2.0 (ref. no. 101-820-200) on a SMRT-cell 8M chip (ref. no. 101-389-001).

All samples were sequenced at the Department of Clinical Genetics at the Amsterdam University Medical Center (AUMC) using a Pacific Biosciences (PacBio) Sequel IIe System with 2 hours of pre-extension time and 30 hours of collection time. Overall, 92 AD genomes were sequenced at a median coverage of 18.2x, with median read-lengths of 14.8 Kbp; 117 centenarian genomes were sequenced at a median coverage of 20.1x, with median read-lengths of 14.7 Kbp.

### PacBio HiFi data processing

After sequencing, raw reads were collected and analyzed through an in-house pipeline (freely available at https://github.com/holstegelab/snakemake_pipeline). Briefly, raw reads first underwent PacBio’s *ccs* algorithm (v6.0.0, https://github.com/PacificBiosciences/ccs) to generate high-fidelity (HiFi) reads with custom parameters (min-passes 0, min-rq 0, keeping kinetics information). Using these parameters, we retained both *HiFi reads* (read-quality >99%, number of passes >3), and lower-quality *non-HiFi reads*, which are typically excluded when running the *ccs* algorithm with default settings. HiFi and non-HiFi reads were then aligned to GRCh38 (patch release 14) reference genomes using *pbmm2* (v1.3.0, https://github.com/PacificBiosciences/pbmm2), with suggested alignment presets for HiFi and non-HiFi reads, generating one BAM-file per SMRT-cell. When available, individual BAM-files from the same sample were merged into a single BAM-file using *samtools*^81^ (v1.13).

We used HiFi-based publicly available whole-genomes to contextualise our findings to global human genetic diversity. Specifically, we downloaded 47 GRCh38-aligned BAM-files from the Human Pangenome Research Consortium (HPRC)^28^ from their respective AWS S3 buckets representing “Year 1” data freeze (https://github.com/human-pangenomics/HPP_Year1_Data_Freeze_v1.0). Long-reads were extracted from the BAM-files using *samtools* and re-aligned to GRCh38 using *minimap2*^82^ (version 2.21-r1071).

### Variant calling and linkage analysis

Structural variants across AD, centenarians, and HPRC genomes were identified using *sniffles2*^83^ (version 2.0.6) with default parameters. Manual inspection of the structural variants in the TMEM106B gene using the UCSC genome browser (https://genome.ucsc.edu/) showed a large deletion overlapping the 3’ UTR nearby known GWAS variants. The deletion exactly overlapped an AluYb8 retrotransposon based on annotations from the UCSC genome browser. This AluYb8-element (chr7:12242091-12242407) along with GWAS variants rs1020004, rs3173615, rs13237518, rs1990622, rs1990621, and rs1990620 were then locally assembled across all AD, centenarian, and HPRC genomes using the *“assemble”* command from *otter* (version 0.1.0, https://github.com/holstegelab/otter) with the “*-F 0.2*” parameter. The locally assembled sequence of the deletion were aligned to DFAM^29^ (release 3.7), confirming the structural variant is indeed an AluYb8 element. The locally assembled sequences were then used to jointly genotype all variants using the “*genotype”* command from *otter* with the *“-e 0.5 -m 10*”, ensuring that only the genotypes of variants with at least 10x of total coverage per region per sample were reported. Pairwise linkage-disequilibrium (LD) was then performed using the “genetics” package (version 1.3.8.1.3) in RStudio (version 2023.06.0+421). GWAS-variants and transcripts in TMEM106B were visualized with *ggbio*^84^ (version 1.42.0) using the *EnsDB.Hsapiens.v86* package^85^, as well as LocusZoom^86^ using the “*Alzheimer_NatGenet2022*” study.

### Comparison of complete *TMEM106B* haplotype sequences

Haplotype-phased *de novo* genome assembly was performed with *hifiasm*^87^ (version 0.16.1-r375) using default parameters on three high-coverage (>30x) centenarian genomes based on PacBio HiFi whole-genome sequencing data generated in this study (Cent1, Cent2, and C3). Haplotype-phase *de novo* genome assemblies from the HPRC “Year 1 Data Freeze” dataset were downloaded^28^ (see “PacBio HiFi data processing“). The centenarian genome assemblies, the CHM13 reference genome (version 2.0)^55^, and HPRC genomes with reduced LD (see section above), along with HG002, were whole-genome aligned to GRCh38, and both single-nucleotide and (large) structural variation were identified with *paftools* (version 2.22-r1101, https://github.com/lh3/minimap2/blob/master/misc/README.md). Variant calls corresponding to the *TMEM106B* locus using GRCh38 coordinates (chr7:12211294-12244382) were merged, and their presence and absence across each haplotype sequence was binarised. Binary clustering was performed with *hclust* in RStudio^88^.

### Profiling methylation in *TMEM106B* locus

Methylation probabilities of CpG sites were calculated using PacBio’s *pg-CpG-tools* (version 2.3.1, https://github.com/PacificBiosciences/pb-CpG-tools) with default parameters along with the “*--modsites-mode reference*” parameter on aligned BAM files to GRCh38. Sequence variation in CpG sites were analysed by identifying corresponding dinucleotide sequences in phased sequences of *TMEM106B*. More specifically, *hifiasm* was used to generate haplotype-phased *de novo* genome assemblies of all AD and centenarian genomes using the default parameters, and whole-genome aligned to GRCh38 with *minimap2* using the “-x asm10” parameter. Only high-quality assemblies—that is, assemblies with exactly two haplotype-phased contigs of at least 100 Kbp and fully spanning *TMEM106B*—were retained. The corresponding positions of each CpG sites reported by *pg-CpG tools* were identified via the whole-genome alignments, and the correpsonding dinucleotide sequences of each haplotype-phased contig were extracted. CpG site coordinates were intersected against regulatory and retro(transposon) element annotations in GRCh38 from the UCSC genome browser. RNA polymerase III and transcription binding recognition sequences within the AluYb8 element were cross-referenced against published annotations from Conti *et al.* 2015^40^. Statistical comparison between carriers of the risk and protective *TMEM106B* haplotypes were done using RStudio^88^.

### Sequence analysis of non-primate genomes

We used the UCSC genome browser (http://genome.ucsc.edu) to identify homologous genes of *TMEM106B* in non-primate genomes, and determine if SINE-retrotransposons are also inserted in their respective 3’ UTR based on available repeat annotations from the UCSC genome browser. Specifically, we used the following reference genomes: mouse (*Mus musculus,* build GRCm39); rat (*Rattus norvegicus*, build RGSC 6.0), pig (*Sus scrofa,* build Sscrofa11.1), dog (*Canis lupus familiaris*, build canFam4) and cat (*Felis catus*, build felCat9).

## Data Availability

All data produced in the present study are available upon reasonable request to the authors.

## Acknowledgements

The authors are grateful to all study participants, their family members, the participating medical staff, general practitioners, pharmacists and all laboratory personnel involved in patient diagnosis, blood collection, blood biobanking, DNA preparation and sequencing. Part of the work in this manuscript was carried out on the Cartesius supercomputer, which is embedded in the Dutch national e-infrastructure with the support of SURF Cooperative. Computing hours were granted to H. H. by the Dutch Research Council (‘100plus’: project# vuh15226, 15318, 17232, and 2020.030; ‘Role of VNTRs in AD’; project# 2022.31, ‘Alzheimer’s Genetics Hub’ project# 2022.38). This work is supported by a VIDI grant from the Dutch Scientific Counsel (#NWO 09150172010083) and a public-private partnership with TU Delft and PacBIo, receiving funding from ZonMW and Health∼Holland, Topsector Life Sciences & Health (PPP-allowance), and by Alzheimer Nederland WE.03-2018-07. H.H., S.L., are recipients of ABOARD, a public-private partnership receiving funding from ZonMW (#73305095007) and Health∼Holland, Topsector Life Sciences & Health (PPP-allowance; #LSHM20106). S.L. is recipient of ZonMW funding (#733050512). H.H. was supported by the Hans und Ilse Breuer Stiftung (2020), Dioraphte 16020404 (2014) and the HorstingStuit Foundation (2018). Acquisition of the PacBio Sequel II long read sequencing machine was supported by the ADORE Foundation (2022).

HH has a collaboration contract with Muna Therapeutics, PacBio, Neurimmune and Alchemab. She serves in the scientific advisory boards of Muna Therapeutics and is an external advisor for Retromer Therapeutics. Research of Alzheimer center Amsterdam is part of the neurodegeneration research program of Amsterdam Neuroscience. Alzheimer Center Amsterdam is supported by Stichting Alzheimer Nederland and Stichting Steun Alzheimercentrum Amsterdam. The clinical database structure was developed with funding from Stichting Dioraphte.

## Author contributions

Conceived the study: HH; Wrote the manuscript: AS, HH; Patient selection: NT, HH, Patient blood collection and sequencing, MG, MH, DD, SW, JK LK; Data management: NT, MH, SvdL, Bioinformatic analysis: AS.

## References

1. Feng, T., Lacrampe, A. & Hu, F. Physiological and pathological functions of TMEM106B: a gene associated with brain aging and multiple brain disorders. Acta Neuropathol. (Berl.) 141, 327–339 (2021).

2. Nicholson, A. M. & Rademakers, R. What we know about TMEM106B in neurodegeneration. Acta Neuropathol. (Berl.) 132, 639–651 (2016).

3. Van Deerlin, V. M. et al. Common variants at 7p21 are associated with frontotemporal lobar degeneration with TDP-43 inclusions. Nat. Genet. 42, 234–239 (2010).

4. Tesi, N., et al. Cognitively Healthy Centenarians are genetically protected against Alzheimer’s disease specifically in immune and endo-lysosomal systems. http://medrxiv.org/lookup/doi/10.1101/2023.05.16.23290049(2023) doi:10.1101/2023.05.16.23290049.

5. Hu, Y. et al. rs1990622 variant associates with Alzheimer’s disease and regulates TMEM106B expression in human brain tissues. BMC Med. 19, 11 (2021).

6. Tropea, T. F. et al. TMEM106B Effect on cognition in Parkinson disease and frontotemporal dementia. Ann. Neurol. 85, 801–811 (2019).

7. Mao, F. et al. TMEM106B modifies TDP-43 pathology in human ALS brain and cell-based models of TDP-43 proteinopathy. Acta Neuropathol. (Berl.) 142, 629–642 (2021).

8. Li, Z. et al. The TMEM106B FTLD-protective variant, rs1990621, is also associated with increased neuronal proportion. Acta Neuropathol. (Berl.) 139, 45–61 (2020).

9. Van Der Zee, J. et al. TMEM106B is associated with frontotemporal lobar degeneration in a clinically diagnosed patient cohort. Brain 134, 808–815 (2011).

10. Gallagher, M. D. et al. A Dementia-Associated Risk Variant near TMEM106B Alters Chromatin Architecture and Gene Expression. Am. J. Hum. Genet. 101, 643–663 (2017).

11. Vass, R. et al. Risk genotypes at TMEM106B are associated with cognitive impairment in amyotrophic lateral sclerosis. Acta Neuropathol. (Berl.) 121, 373–380 (2011).

12. Bellenguez, C. et al. New insights into the genetic etiology of Alzheimer’s disease and related dementias. Nat. Genet. 54, 412–436 (2022).

13. Rhinn, H., Tatton, N., McCaughey, S., Kurnellas, M. & Rosenthal, A. Progranulin as a therapeutic target in neurodegenerative diseases. Trends Pharmacol. Sci. 43, 641–652 (2022).

14. Cabron, A.-S. et al. Lack of a protective effect of the Tmem106b “protective SNP” in the Grn knockout mouse model for frontotemporal lobar degeneration. Acta Neuropathol. Commun. 11, 21 (2023).

15. Harding, S. R. et al. The TMEM106B risk allele is associated with lower cortical volumes in a clinically diagnosed frontotemporal dementia cohort. J. Neurol. Neurosurg. Psychiatry 88, 997–998 (2017).

16. Chen-Plotkin, A. S., et al. *TMEM106B*, the Risk Gene for Frontotemporal Dementia, Is Regulated by the microRNA-132/212 Cluster and Affects Progranulin Pathways. J. Neurosci. 32, 11213–11227 (2012).

17. Nicholson, A. M. et al. TMEM106B p.T185S regulates TMEM106B protein levels: implications for frontotemporal dementia. J. Neurochem. 126, 781–791 (2013).

18. T Vicente, C., et al. C-terminal TMEM106B fragments in human brain correlate with disease-associated *TMEM106B* haplotypes. Brain awad133 (2023) doi:10.1093/brain/awad133.

19. Lang, C. M. et al. Membrane Orientation and Subcellular Localization of Transmembrane Protein 106B (TMEM106B), a Major Risk Factor for Frontotemporal Lobar Degeneration. J. Biol. Chem. 287, 19355–19365 (2012).

20. Schweighauser, M. et al. Age-dependent formation of TMEM106B amyloid filaments in human brains. Nature 605, 310–314 (2022).

21. Jiang, Y. X. et al. Amyloid fibrils in FTLD-TDP are composed of TMEM106B and not TDP-43. Nature 605, 304–309 (2022).

22. Chang, A. et al. Homotypic fibrillization of TMEM106B across diverse neurodegenerative diseases. Cell 185, 1346–1355.e15 (2022).

23. Fan, Y. et al. Generic amyloid fibrillation of TMEM106B in patient with Parkinson’s disease dementia and normal elders. Cell Res. 32, 585–588 (2022).

24. Fan, Y. et al. Newly identified transmembrane protein 106B amyloid fibrils in the human brain: pathogens or by-products? Ageing Neurodegener. Dis. 3, 4 (2023).

25. Ren, Y. et al. TMEM106B haplotypes have distinct gene expression patterns in aged brain. Mol. Neurodegener. 13, 35 (2018).

26. Yang, H.-S. et al. Genetics of Gene Expression in the Aging Human Brain Reveal TDP-43 Proteinopathy Pathophysiology. Neuron 107, 496–508.e6 (2020).

27. Chemparathy, A., et al. *A 3’ UTR Deletion Is a Leading Candidate Causal Variant at the* TMEM106B *Locus Reducing Risk for FTLD-TDP*. http://medrxiv.org/lookup/doi/10.1101/2023.07.06.23292312(2023) doi:10.1101/2023.07.06.23292312.

28. Wang, T. et al. The Human Pangenome Project: a global resource to map genomic diversity. Nature 604, 437–446 (2022).

29. Storer, J., Hubley, R., Rosen, J., Wheeler, T. J. & Smit, A. F. The Dfam community resource of transposable element families, sequence models, and genome annotations. Mob. DNA 12, 2 (2021).

30. Deininger, P. Alu elements: know the SINEs. Genome Biol. 12, 236 (2011).

31. Chen, L.-L. & Yang, L. ALU ternative Regulation for Gene Expression. Trends Cell Biol. 27, 480–490 (2017).

32. Solyom, S. & Kazazian, H. H. Mobile elements in the human genome: implications for disease. Genome Med. 4, 12 (2012).

33. Feusier, J. et al. Discovery of rare, diagnostic AluYb8/9 elements in diverse human populations. Mob. DNA 8, 9 (2017).

34. Carter, A. B. et al. Genome-wide analysis of the human Alu Yb-lineage. Hum. Genomics 1, 167 (2004).

35. Morera, A. A., Ahmed, N. S. & Schwartz, J. C. TDP-43 regulates transcription at protein-coding genes and Alu retrotransposons. Biochim. Biophys. Acta BBA - Gene Regul. Mech. 1862, 194434 (2019).

36. Almeida, M. V., Vernaz, G., Putman, A. L. K. & Miska, E. A. Taming transposable elements in vertebrates: from epigenetic silencing to domestication. Trends Genet. 38, 529–553 (2022).

37. Smit, A. F. A. & Riggs, A. D. MIRs are classic, tRNA-derived SINEs that amplified before the mammalian radiation. Nucleic Acids Res. 23, 98–102 (1995).

38. Goodier, J. L. Restricting retrotransposons: a review. Mob. DNA 7, 16 (2016).

39. Carlevaro-Fita, J. et al. Ancient exapted transposable elements promote nuclear enrichment of human long noncoding RNAs. Genome Res. 29, 208–222 (2019).

40. Conti, A. et al. Identification of RNA polymerase III-transcribed Alu loci by computational screening of RNA-Seq data. Nucleic Acids Res. 43, 817–835 (2015).

41. Kochanek, S., Renz, D. & Doerfler, W. Transcriptional silencing of human Alu sequences and inhibition of protein binding in the box B regulatory elements by 5ʹ-CG-3ʹ methylation. FEBS Lett. 360, 115–120 (1995).

42. Jordà, M. et al. The epigenetic landscape of *Alu* repeats delineates the structural and functional genomic architecture of colon cancer cells. Genome Res. 27, 118–132 (2017).

43. Grechishnikova, D. & Poptsova, M. Conserved 3ʹ UTR stem-loop structure in L1 and Alu transposons in human genome: possible role in retrotransposition. BMC Genomics 17, 992 (2016).

44. Baar, T. et al. RNA transcription and degradation of Alu retrotransposons depends on sequence features and evolutionary history. G3 GenesGenomesGenetics 12, jkac054 (2022).

45. Li, W., Jin, Y., Prazak, L., Hammell, M. & Dubnau, J. Transposable Elements in TDP-43-Mediated Neurodegenerative Disorders. PLoS ONE 7, e44099 (2012).

46. Tam, O. H. et al. Postmortem Cortex Samples Identify Distinct Molecular Subtypes of ALS: Retrotransposon Activation, Oxidative Stress, and Activated Glia. Cell Rep. 29, 1164–1177.e5 (2019).

47. Koike, Y. et al. Age-related demethylation of the TDP-43 autoregulatory region in the human motor cortex. *Commun*. Biol. 4, 1107 (2021).

48. Lucas, B. A. et al. Evidence for convergent evolution of SINE-directed Staufen-mediated mRNA decay. Proc. Natl. Acad. Sci. 115, 968–973 (2018).

49. Maquat, L. E. Short interspersed nuclear element (SINE)-mediated post-transcriptional effects on human and mouse gene expression: SINE-UP for active duty. Philos. Trans. R. Soc. B Biol. Sci. 375, 20190344 (2020).

50. Mouse Genome Sequencing Consortium. Initial sequencing and comparative analysis of the mouse genome. Nature 420, 520–562 (2002).

51. Ferraj, A. et al. Resolution of structural variation in diverse mouse genomes reveals chromatin remodeling due to transposable elements. Cell Genomics 3, 100291 (2023).

52. Funkhouser, S. A. et al. Evidence for transcriptome-wide RNA editing among Sus scrofa PRE-1 SINE elements. BMC Genomics 18, 360 (2017).

53. Fang, X. et al. The sequence and analysis of a Chinese pig genome. GigaScience 1, 16 (2012).

54. Wang, W. & Kirkness, E. F. Short interspersed elements (SINEs) are a major source of canine genomic diversity. Genome Res. 15, 1798–1808 (2005).

55. Nurk, S. et al. The complete sequence of a human genome. Science 376, 44–53 (2022).

56. Deininger, P. L. & Batzer, M. A. Alu Repeats and Human Disease. Mol. Genet. Metab. 67, 183–193 (1999).

57. Gussakovsky, D. & McKenna, S. A. Alu RNA and their roles in human disease states. RNA Biol. 18, 574–585 (2021).

58. Ciccarone, F., Tagliatesta, S., Caiafa, P. & Zampieri, M. DNA methylation dynamics in aging: how far are we from understanding the mechanisms? Mech. Ageing Dev. 174, 3–17 (2018).

59. Lee, J.-H., Kim, E. W., Croteau, D. L. & Bohr, V. A. Heterochromatin: an epigenetic point of view in aging. Exp. Mol. Med. 52, 1466–1474 (2020).

60. Mustafina, O. E. The possible roles of human Alu elements in aging. Front. Genet. 4, (2013).

61. Larsen, P. A. et al. The *Alu* neurodegeneration hypothesis: A primate-specific mechanism for neuronal transcription noise, mitochondrial dysfunction, and manifestation of neurodegenerative disease. Alzheimers Dement. 13, 828–838 (2017).

62. De Cecco, M. et al. Transposable elements become active and mobile in the genomes of aging mammalian somatic tissues. Aging 5, 867–883 (2013).

63. Zhang, X.-O., Gingeras, T. R. & Weng, Z. Genome-wide analysis of polymerase III– transcribed *Alu* elements suggests cell-type–specific enhancer function. Genome Res. 29, 1402–1414 (2019).

64. Gao, Z., Herrera-Carrillo, E. & Berkhout, B. RNA Polymerase II Activity of Type 3 Pol III Promoters. Mol. Ther. - Nucleic Acids 12, 135–145 (2018).

65. Daskalova, E., Baev, V., Rusinov, V. & Minkov, I. 3’UTR-located *ALU* Elements: Donors of Potetial miRNA Target Sites and Mediators of Network miRNA-based Regulatory Interactions. Evol. Bioinforma. 2, 117693430600200 (2006).

66. Pandey, R. et al. Alu-miRNA interactions modulate transcript isoform diversity in stress response and reveal signatures of positive selection. Sci. Rep. 6, 32348 (2016).

67. Gong, C. & Maquat, L. E. lncRNAs transactivate STAU1-mediated mRNA decay by duplexing with 3ʹ UTRs via Alu elements. Nature 470, 284–288 (2011).

68. Park, E. & Maquat, L. E. Staufen-mediated mRNA decay: Staufen-mediated mRNA decay. Wiley Interdiscip. Rev. RNA 4, 423–435 (2013).

69. Liang, L. et al. Complementary Alu sequences mediate enhancer–promoter selectivity. Nature 619, 868–875 (2023).

70. Alharbi, A. B., Schmitz, U., Bailey, C. G. & Rasko, J. E. J. CTCF as a regulator of alternative splicing: new tricks for an old player. Nucleic Acids Res. 49, 7825–7838 (2021).

71. Fiesel, F. C. et al. Knockdown of transactive response DNA-binding protein (TDP-43) downregulates histone deacetylase 6. EMBO J. 29, 209–221 (2010).

72. Seddighi, S., et al. *Mis-spliced transcripts generate* de novo *proteins in TDP-43-related ALS/FTD*. http://biorxiv.org/lookup/doi/10.1101/2023.01.23.525149(2023) doi:10.1101/2023.01.23.525149.

73. Lee, J. Y. et al. The major TMEM106B dementia risk allele affects TMEM106B protein levels, fibril formation, and myelin lipid homeostasis in the ageing human hippocampus. Mol. Neurodegener. 18, 63 (2023).

74. Tian, J. et al. CancerSplicingQTL: a database for genome-wide identification of splicing QTLs in human cancer. Nucleic Acids Res. 47, D909–D916 (2019).

75. Baggen, J. et al. TMEM106B is a receptor mediating ACE2-independent SARS-CoV-2 cell entry. Cell 186, 3427–3442.e22 (2023).

76. Baggen, J. et al. Genome-wide CRISPR screening identifies TMEM106B as a proviral host factor for SARS-CoV-2. Nat. Genet. 53, 435–444 (2021).

77. Lake, J. et al. Multi-ancestry meta-analysis and fine-mapping in Alzheimer’s disease. Mol. Psychiatry (2023) doi:10.1038/s41380-023-02089-w.

78. Sherva, R. et al. African ancestry GWAS of dementia in a large military cohort identifies significant risk loci. Mol. Psychiatry 28, 1293–1302 (2023).

79. Van Der Flier, W. M. & Scheltens, P. Amsterdam Dementia Cohort: Performing Research to Optimize Care. J. Alzheimers Dis. 62, 1091–1111 (2018).

80. Holstege, H. et al. The 100-plus Study of cognitively healthy centenarians: rationale, design and cohort description. Eur. J. Epidemiol. 33, 1229–1249 (2018).

81. Li, H. et al. The Sequence Alignment/Map format and SAMtools. Bioinformatics 25, 2078–2079 (2009).

82. Li, H. Minimap2: pairwise alignment for nucleotide sequences. Bioinformatics 34, 3094–3100 (2018).

83. Smolka, M., et al. Comprehensive Structural Variant Detection: From Mosaic to Population-Level. http://biorxiv.org/lookup/doi/10.1101/2022.04.04.487055(2022) doi:10.1101/2022.04.04.487055.

84. Yin, T., Cook, D. & Lawrence, M. ggbio: an R package for extending the grammar of graphics for genomic data. Genome Biol. 13, R77 (2012).

85. Rainer, J. EnsDb.Hsapiens.v86. (2017) doi:10.18129/B9.BIOC.ENSDB.HSAPIENS.V86.

86. Boughton, A. P. et al. LocusZoom.js: interactive and embeddable visualization of genetic association study results. Bioinformatics 37, 3017–3018 (2021).

87. Cheng, H., Concepcion, G. T., Feng, X., Zhang, H. & Li, H. Haplotype-resolved de novo assembly using phased assembly graphs with hifiasm. Nat. Methods 18, 170–175 (2021).

88. R Core Team. R: A Language and Environment for Statistical Computing. (R Foundation for Statistical Computing, 2021).

